# The combined effect of lifestyle factors and polygenic scores on age at onset in Parkinson’s disease

**DOI:** 10.1101/2023.08.25.23294466

**Authors:** Carolin Gabbert, Leonie Blöbaum, Theresa Lüth, Inke R. König, Amke Caliebe, Sebastian Koch, Björn-Hergen Laabs, Christine Klein, Joanne Trinh

## Abstract

**Objective:** To investigate the association between a Parkinson’s disease (PD)-specific polygenic score (PGS) and protective lifestyle factors on age at onset (AAO) in PD.

**Methods:** We included data from 4375 patients with idiopathic PD, 167 patients with *GBA1*-PD, and 3091 healthy controls of European ancestry from AMP-PD, PPMI, and Fox Insight cohorts. The PGS was calculated based on a previously proposed composition of 1805 variants. The association between PGS and lifestyle factors (i.e., coffee, tobacco, and aspirin) on AAO was assessed with linear and Cox proportional hazards models.

**Results:** The PGS showed a negative association with AAO (β=-1.07, p=6x10^-7^). The use of one, two, or three of the protective lifestyle factors showed a reduction in the hazard ratio by 21% (p=0.0001), 45% (p<2x10^-16^), and 55% (p<2x10^-16^), respectively, compared to no use. An additive effect of aspirin (β=7.61, p=8x10^-7^) and PGS (β=-1.63, p=0.0112) was found for AAO without an interaction (p=0.9789) in the linear regressions, and similar effects were seen for tobacco. Aspirin is shown to be a better predictor of AAO (R^2^=0.1740) compared to coffee and tobacco use (R^2^=0.0243, R^2^=0.0295) or the PGS (R^2^=0.0141). In contrast, no association between aspirin and AAO was found in *GBA1*-PD (p>0.05).

**Interpretation:** In our cohort, coffee, tobacco, aspirin, and PGS are independent predictors of PD AAO. Additionally, lifestyle factors seem to have a greater influence on AAO than common genetic risk variants with aspirin presenting the largest effect. External validation of our findings is needed.

## Introduction

Parkinson’s disease (PD) is a complex neurodegenerative disorder. Besides monogenic forms of PD that explain about 5% of PD cases^1^, GWAS studies have shown that idiopathic PD is highly polygenic^2,3^. The largest meta-GWAS of PD to date identified 90 independent risk loci across 78 genomic regions that explained between 16% to 36% of the heritable risk of PD^2^. That study additionally determined the proportion of SNP-based heritability explained by their PD GWAS and found their 1805 variant polygenic score (PGS) to explain about 26% of PD heritability^2^. The calculation of PGSs provides the opportunity to summarize the effect of the heritable risk to develop the disease on the individual level. Several studies already evaluated the association of PGSs for PD and affection status, age at onset (AAO), or PD-related symptoms^4–12^.

In addition to common genetic risk factors, environmental and lifestyle factors have consistently shown an association with PD susceptibility. In several studies, a protective effect of the use of caffeine, tobacco, or aspirin for PD risk has already been observed^13–19^. Additionally, interactions between genetic modifiers and lifestyle factors can affect PD risk. Gene-environment interactions have been shown between the genetic assembly and a patient’s lifestyle. Thus far, there are known interactions between *GRIN2A*, *ADORA2A*, and *CYP1A2* and coffee^20, 21^ as well as between *RXRA*, *SLC17A6*, and *HLA-DRB1* and smoking^22,23^. In contrast, studies investigating the effect of environmental and lifestyle factors or gene-environment interactions on PD AAO are only sparse. While some studies found a protective effect of coffee and smoking on PD AAO^24–32^, literature on the effect of aspirin on AAO is lacking^31^. It also remains unclear how gene-environment interactions or a genetic predisposition to PD risk together with the presence of certain lifestyle factors influences the AAO in PD.

Herein, we examine AAO associations of the PGS and the combined effect of coffee drinking, tobacco use, and aspirin intake in PD. We investigate whether coffee drinking, tobacco use, and aspirin intake are positively associated with the AAO in PD and if these lifestyle factors further have an additive or interactive effect with respect to the PGS on PD AAO.

## Methods

### Study demographics

Three datasets containing genetic, environmental, and lifestyle data from the Accelerating Medicine Partnership Parkinson’s Disease Knowledge Platform (AMP-PD), the Parkinson’s Progression Markers Initiative (PPMI), and the Fox Insight cohort were included in this study. Patients and healthy controls from PPMI are included in the AMP-PD cohort for genome sequencing and more detailed lifestyle data was documented as part of the PPMI cohort. In total, 7633 unrelated participants were included in our study: 4375 patients with PD, without a known genetic cause of PD, 167 patients with variants in *GBA1,* which harbor some of the strongest genetic risk variants in PD, and 3091 healthy controls (Table 1). In this study group, the mean AAO of patients with PD without a known genetic cause of PD was 60.5 years (standard deviation, SD = ± 9.7 years, range: 19.3 to 89.1 years) and the mean age at examination (AAE) was 64.7 years (SD = ± 9.0 years, range: 33.0 to 91.5 years). Of the patients with PD, 2482 (56.7%) were men and 1892 (43.2%) were women. Of the 4375 patients with PD, 1987 were from the AMP-PD cohort, of which 387 were from the PPMI subgroup of AMP-PD, and 2378 were from the Fox Insight cohort. The group of patients with *GBA1*-PD, who carried one of the GBA1 variants p.R535H (NM_000157.4, c.1604G>A), p.N409S (NM_000157.4, c.1226A>G), and p.E365K (NM_000157.4, c.1093G>A) consisted of 167 patients with a mean AAO of 61.3 years (SD = ± 10.1 years, range: 28.5 to 83.9 years) and a mean AAE of 65.2 years (SD = ± 9.4 years, range: 32.0 to 86.1 years). Of these, 78 (46.7%) were men and 81 (48.5%) were women, while gender information was missing for eight patients. The group of healthy controls consisted of 3091 participants with a mean AAE of 69.9 years (SD = ± 13.0 years, range: 16.0 to 90.0 years). While 1495 (48.4%) of the controls were men, 1595 (51.6%) were women. All participants included in this study were of white European ancestry.

**Table 1.**
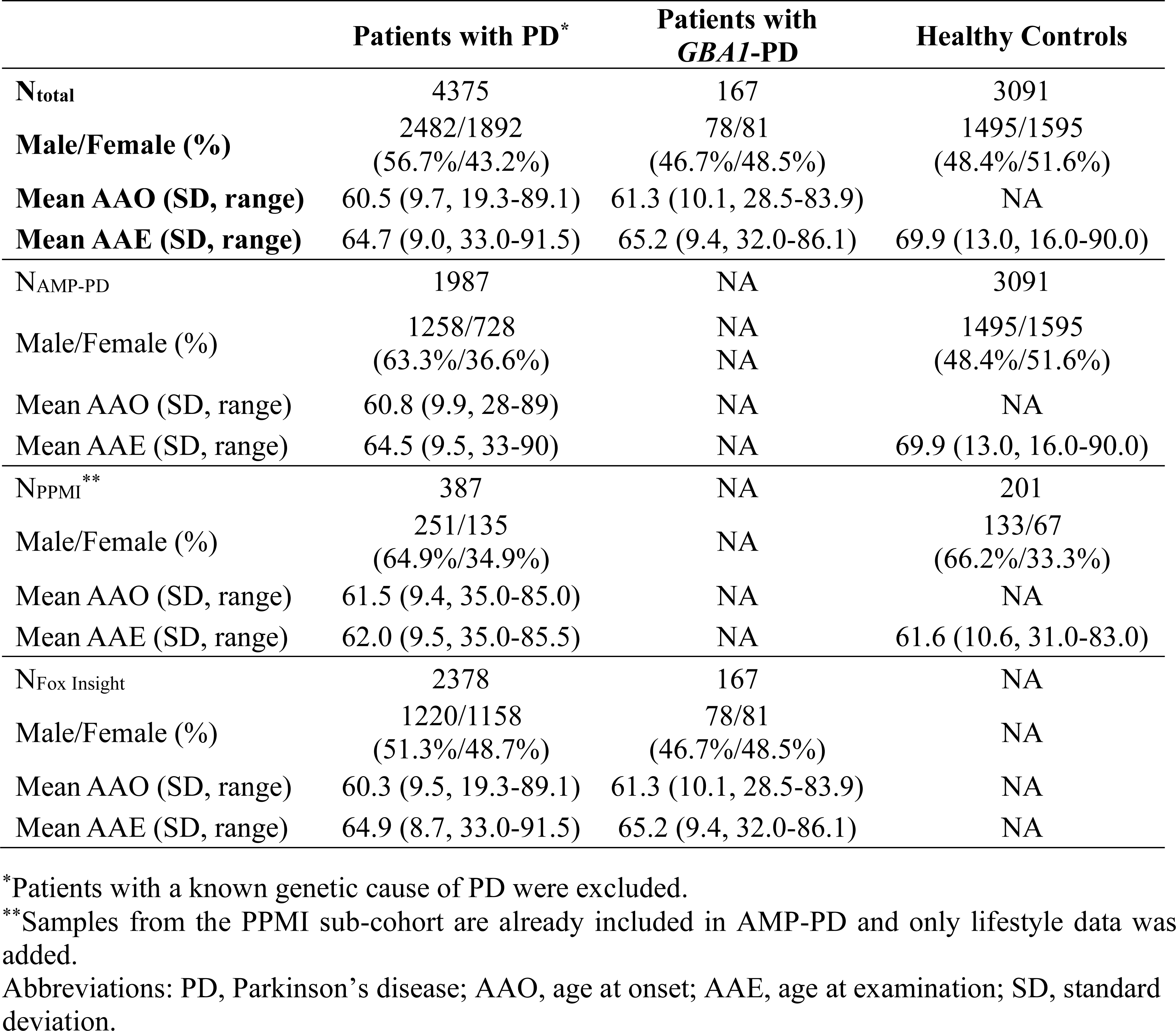
Demographics of the study group.

|  | Patients with PD* | Patients with *GBA1*-PD | Healthy Controls |
| --- | --- | --- | --- |
| **N<sub>total</sub>** | 4375 | 167 | 3091 |
| **Male/Female (%)** | 2482/1892<br>(56.7%/43.2%) | 78/81<br>(46.7%/48.5%) | 1495/1595<br>(48.4%/51.6%) |
| **Mean AAO (SD, range)** | 60.5 (9.7, 19.3-89.1) | 61.3 (10.1, 28.5-83.9) | NA |
| **Mean AAE (SD, range)** | 64.7 (9.0, 33.0-91.5) | 65.2 (9.4, 32.0-86.1) | 69.9 (13.0, 16.0-90.0) |
| **N<sub>AMP-PD</sub>** | 1987 | NA | 3091 |
| **Male/Female (%)** | 1258/728<br>(63.3%/36.6%) | NA<br>NA | 1495/1595<br>(48.4%/51.6%) |
| **Mean AAO (SD, range)** | 60.8 (9.9, 28-89) | NA | NA |
| **Mean AAE (SD, range)** | 64.5 (9.5, 33-90) | NA | 69.9 (13.0, 16.0-90.0) |
| **N<sub>PPMI</sub>\*\*** | 387 | NA | 201 |
| **Male/Female (%)** | 251/135<br>(64.9%/34.9%) | NA | 133/67<br>(66.2%/33.3%) |
| **Mean AAO (SD, range)** | 61.5 (9.4, 35.0-85.0) | NA | NA |
| **Mean AAE (SD, range)** | 62.0 (9.5, 35.0-85.5) | NA | 61.6 (10.6, 31.0-83.0) |
| **N<sub>Fox Insight</sub>** | 2378 | 167 | NA |
| **Male/Female (%)** | 1220/1158<br>(51.3%/48.7%) | 78/81<br>(46.7%/48.5%) | NA |
| **Mean AAO (SD, range)** | 60.3 (9.5, 19.3-89.1) | 61.3 (10.1, 28.5-83.9) | NA |
| **Mean AAE (SD, range)** | 64.9 (8.7, 33.0-91.5) | 65.2 (9.4, 32.0-86.1) | NA |

\*Patients with a known genetic cause of PD were excluded.
\*\*Samples from the PPMI sub-cohort are already included in AMP-PD and only lifestyle data was added.
Abbreviations: PD, Parkinson's disease; AAO, age at onset; AAE, age at examination; SD, standard deviation.

### Genetic data and polygenic score estimate

AMP-PD genetic data contained whole-genome sequencing (WGS) data from six unified cohorts (BioFIND, HBS, PDBP, PPMI, SURE-PD3, LBD)^33^. All samples of the AMP-PD dataset were processed by the TOPMed Freeze 9 Variant Calling Pipeline for joint genotyping^33^. The genetic dataset from AMP-PD was stored in a binary PLINK format^34^. The dataset was filtered using PLINK 1.9 according to standard quality control filtering steps, excluding SNPs with a minor allele frequency <0.01, a missingness per sample >0.02, a missingness per SNP >0.05, and that failed Hardy-Weinberg equilibrium at a threshold of 1×10^-50^.

For the PGS calculation, a previously proposed composition of 1805 variants associated with PD risk^2, 4^ was used together with the reference alleles and effect sizes. In our study sample, 1725 of the PGS SNPs were included in the AMP-PD data, with additional 13 SNPs that were represented by proxy SNPs. Proxy SNPs were evaluated with *SNiPA*^35^ and had to be in a linkage disequilibrium of >0.98 with the SNP of interest. In total 1738 SNPs were used for the PGS calculation with the PLINK *score* function. The PGS values were subsequently standardized by subtraction of the mean and division by the standard deviation of the PGS among controls^4^. This standardized PGS was used for all further analyses. Density plots were created with the base-R function *density* and receiver operating characteristic (ROC) curves and corresponding areas under the curve (AUCs) were calculated with the R package *pROC* (R version 4.3.0)^36, 37^.

In order to perform a principal component analysis (PCA), the unfiltered genetic dataset from AMP-PD was pruned based on the linkage disequilibrium (LD) using an R^2^ threshold of 0.3 with the PLINK 1.9 *indep-pairwise* function. Subsequently, the dataset was filtered for a minor allele frequency >0.3 and a genotyping rate >0.99. The PCA was performed using PLINK 1.9 *pca* function.

### Lifestyle and environmental data

Available environmental and lifestyle data were harmonized across the unified cohorts. PD Risk Factor Questionnaire (PD-RFQ) for coffee, tobacco, and aspirin or similar assessments were mainly used^38^. In this study group, patients were classified as coffee consumers if they regularly drank caffeinated coffee at least once per week over a period of at least six months. Patients were classified as tobacco users if they have ever used tobacco, or when available, if they smoked more than 100 cigarettes in their lifetime or if they smoked at least one cigarette per day over a minimum period of six months or if they used smokeless tobacco at least once per day for more than six months. Lastly, patients were classified as aspirin users if they took at least two pills per week over a minimum of six months.

Duration of caffeine consumption, smoking, and aspirin intake were estimated according to the age the patients started using either substance subtracted from the age at termination. If the patients terminated the consumption after their AAO, the age the patients started was subtracted from their AAO. Periods, where the patients stopped regularly consuming, were subtracted from the overall duration.

Coffee drinking dosage was defined as the average number of cups of coffee per day the patients drank within the drinking duration time. Smoking dosage was estimated as cigarettes smoked per day within the smoking duration time. Aspirin dosage was defined as pills per week the patients took within the aspirin intake duration time. The number of cups of coffee for non-drinkers, cigarettes for non-smokers, and pills per week for aspirin non-users was set to zero.

### Statistical analysis

Statistical analyses were performed using R v4.3.0^39^. Multiple linear regression models were used to evaluate the association between AAO, PGS, and lifestyle factors in patients with PD. All linear regression models were validated by evaluating diagnostic plots (Residuals vs Fitted, Q-Q Residuals, Scale-Location, and Residuals vs Leverage) and outliers were removed if applicable. In the linear models, AAO was used as the dependent variable and the PGS and/or the lifestyle factors as the independent variables. Estimates (β), standard errors (SE) and p-values were reported. To adjust for potential confounders, sex and the first two principal components (PCs) were included as covariables in the models. Reported p-values were not corrected for multiple testing because they did not follow an “a priori” hypothesis and results were exploratory. Lifestyle factors were handled in three different ways in the regression models: 1) binary (ever-never/yes-no indication), 2) dosage as a continuous variable, and 3) duration as a continuous variable. In a second set of regression models, data from patients with PD and healthy controls was used to estimate Cox proportional hazards models. Here we modeled AAO from the cumulative number of lifestyle factors used (R package *survival*) and we used AAO for patients with PD and AAE with censoring for healthy controls. The total number of the three lifestyle factors coffee, tobacco, and aspirin the participants used were included as the numbers zero to three. The sex and the study site were additionally included as covariables since genetic data and thus genetic PCs were not available for all participants. Survival plots and forest plots were generated to visualize the Cox proportional hazards model using the *ggsurvplot* (R package *survminer*) and *forest_model* function (R package *forestmodel*). Regression coefficients, hazards ratios (HR), 95% confidence intervals (95% CI) and p-values were reported. To compare the model accuracies of different linear regression models, the adjusted deviance-based R^2^ was calculated using the *adjR2* function (R package *glmtoolbox*). To compare the AAO ranges in patients with PD with respect to their PGS, the patients were stratified into quartiles according to their PGS and the difference in AAO between groups was calculated. In addition, to compare the effect sizes of PGS and lifestyle factors on AAO, the PGS was categorized into “low PGS” and “high PGS” according to the median PGS and participants were stratified into the subgroups that either used no protective lifestyle factor or that used all three lifestyle factors.

## Results

### Relationship between PGS and AAO

First, to validate the PD-specific PGS in this study group, the PGS values of patients with PD and healthy controls were assessed. In a case-control comparison, the AUC for the ROC curves of the standardized PGS was 0.67, which was comparable to the AUC obtained in the original study^2^.

To analyze the association between the PGS and AAO in patients with PD, a linear regression model including sex and the first two PCs as covariates was used. The PGS showed a negative association with AAO (β=-1.07, SE=0.21, p=6x10^-7^). Thus, if the PGS is increased by one standard deviation (SD), the estimated AAO is approximately one year earlier in patients with PD.

We also assessed the AAO ranges in patients with PD by stratifying and comparing the first and last PGS quartiles. Patients with PD in the first PGS quartile had a median AAO of 63 years (range: 31-85 years), while PD patients in the last PGS quartile had a median AAO of 61 years (range: 34-83 years), showing a difference in the median AAO of two years in these two groups.

### Relationship between lifestyle factors and AAO

We replicated our previous findings from the Fox Insight cohort^31^ in the AMP-PD/PPMI cohort. In the linear regression model, coffee drinking duration was positively associated with AAO (β=0.18, SE=0.04, p=4x10^-5^). In addition, tobacco use showed a positive association with AAO (β=3.26, SE=0.50, p=1x10^-10^). We also observed positive associations between aspirin use (β=7.30, SE=1.49, p=3x10^-6^), aspirin dosage (β=0.79, SE=0.21, p=0.0003), and aspirin duration (β=0.54, SE=0.15, p=0.0005) and the AAO. To investigate the additive effect of the three lifestyle factors, we coded them by the cumulative number of factors the patients consumed. In this linear regression model, the use of two (β=4.82, SE=2.38, p=0.0446) or three (β=7.63, SE=2.80, p=0.0073) protective lifestyle factors showed an association with AAO, while the use of one lifestyle factor (β=0.33, SE=2.28, p=0.8859) was not associated with AAO. Interestingly, when including all three factors separately in the same model to predict AAO, aspirin was still associated with AAO (β=6.97, SE=1.53, p=1x10^-5^) and the other associations diminished. Although all three protective factors are associated with AAO, aspirin is shown to be a better predictor of AAO when only one lifestyle factor was included in the model (R^2^=0.1740; aspirin (yes/no), sex, PC1, and PC2 in the model) compared to coffee and tobacco use (R^2^=0.0243, R^2^=0.0295; coffee (yes/no) or tobacco (ever/never), sex, PC1, and PC2 in the model). In a combined analysis of individuals from both AMP-PD/PPMI and Fox Insight using Cox proportional hazards models on the AAO of patients with PD while including the AAE of healthy controls, we first included the lifestyle factors as separate factors. Coffee (HR=0.75, 95% CI=0.68-0.82, p=8x10^-10^), tobacco (HR=0.78, 95% CI=0.72-0.85, p=7x10^-9^), and aspirin (HR=0.66, 95% CI=0.60-0.71, p<2x10^-16^) showed a reduction in the hazard ratio compared to no use by 25%, 22%, and 34%, respectively. To investigate the potential additive effect between all three lifestyle factors, they were coded by the cumulative number of lifestyle factors the participants consumed as above. In the Cox proportional hazards model, the use of one (HR=0.79, 95% CI=0.70-0.89, p=0.0001), two (HR=0.55, 95% CI=0.49-0.62, p<2x10^-16^), or three (HR=0.45, 95% CI=0.38-0.53, p<2x10^-16^) of the selected lifestyle factors showed a reduction in the hazard ratio compared to the use of none of these lifestyle factors by 21%, 45%, and 55%, respectively, indicating a later AAO when using more lifestyle factors (Table 2, Figure 1, Figure 2).

**Figure 1.**
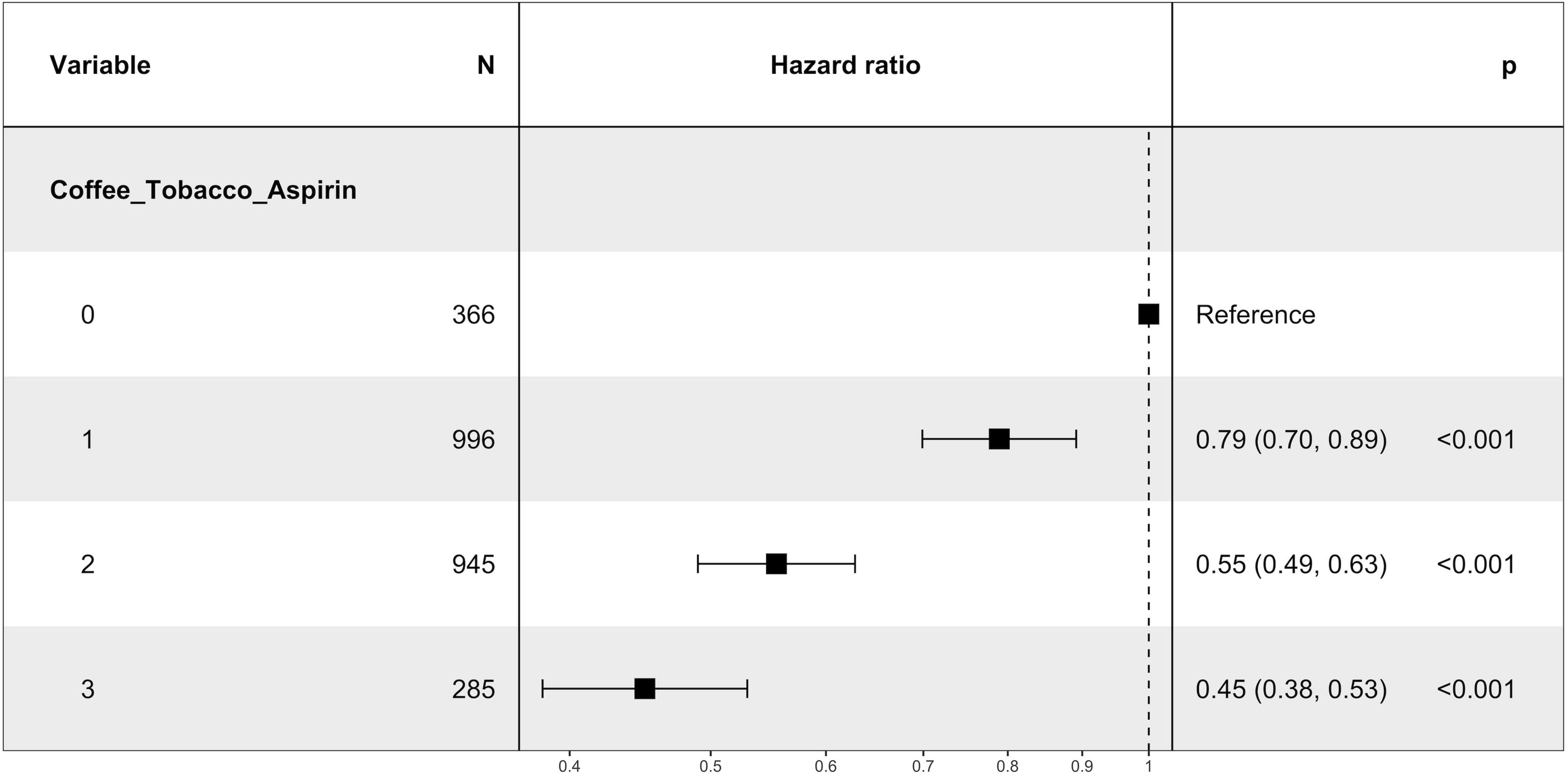
Additive effects of the lifestyle factors coffee drinking, tobacco use, and aspirin intake on the AAO of PD patients, while censoring with the AAE of healthy controls. The different curves describe the cumulative number (0-3) of protective lifestyle factors (coffee drinking, tobacco use, and aspirin intake) the participants used. A Cox proportional hazards model was used to investigate the difference in AAO with respect to the number of protective lifestyle factors used while censoring with the AAE of healthy controls. The sex and study site were additionally included as covariates (→ coxph(formula = Surv(AAO/AAE, Diagnosis) ∼ Coffee/Tobacco/Aspirin + Sex + Study, data = data)).

**Figure 2.**
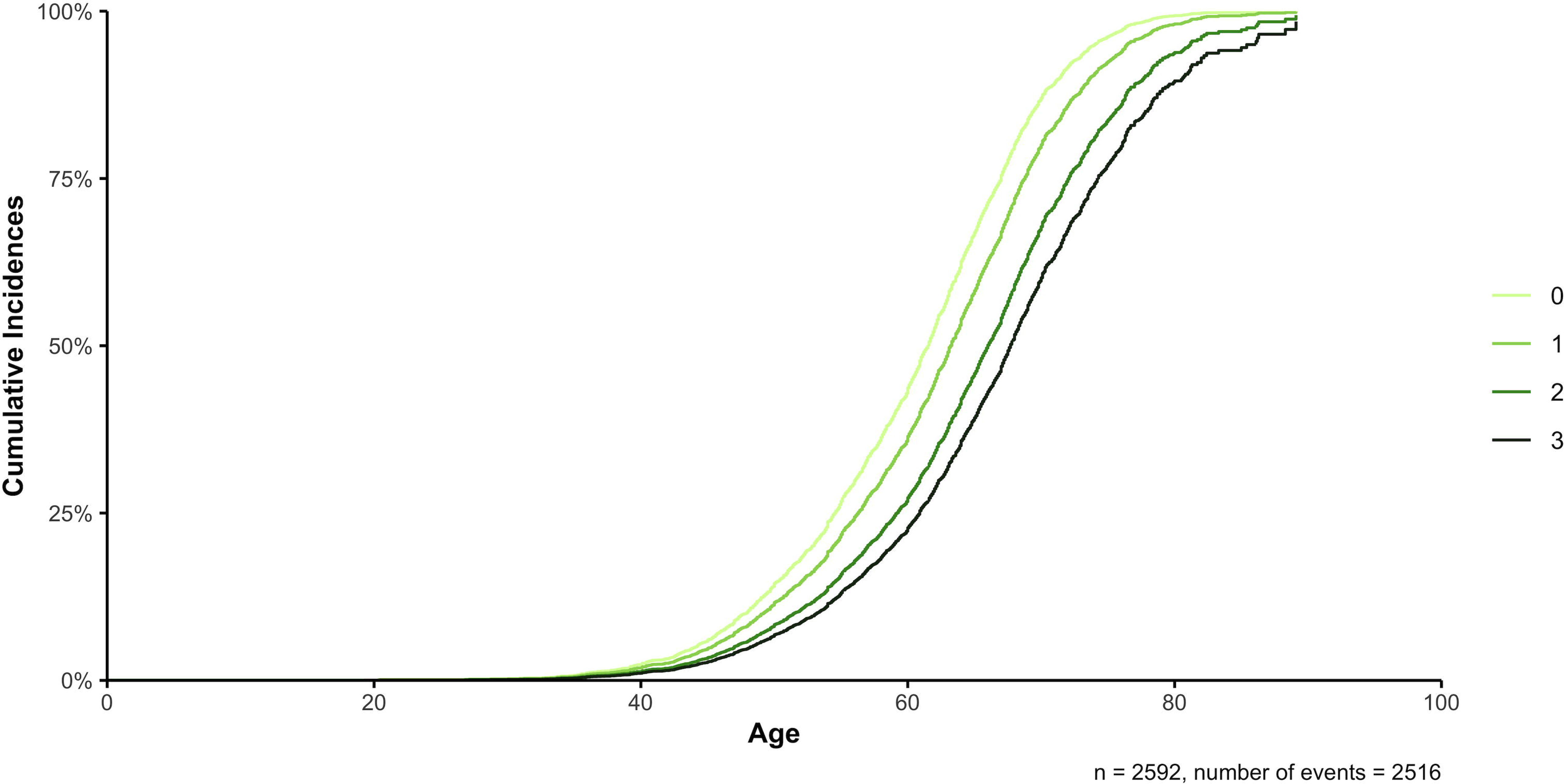
Plot of the Cox proportional hazards model to investigate the additive effects between the use of the lifestyle factors coffee drinking, tobacco use, and aspirin intake (cumulative number (0-3)) on the AAO of PD patients, while censoring with the AAE of healthy controls. The sex and study site were additionally included as covariables but are not displayed. (→ coxph(formula = Surv(AAO/AAE, Diagnosis) ∼ Coffee/Tobacco/Aspirin + Sex + Study, data = data)).

**Table 2.** Cox proportional hazards model to investigate the additive effects between the use of the lifestyle factors coffee drinking, tobacco use, and aspirin intake (cumulative number (0-3)) on the AAO of PD, while censoring with the AAE of healthy controls.

|  | **Regression**<br>**coefficient** | **Hazard Ratio (95% CI)** | **p-value** |  |
| --- | --- | --- | --- | --- |
| **N=2592, N events=2516** |  |  |  |  |
| Coffee/Tobacco/Aspirin (1) | -0.2366 | 0.7893 (0.6989, 0.8914) | 0.0001 | * |
| Coffee/Tobacco/Aspirin (2) | -0.5891 | 0.5548 (0.4900, 0.6283) | <2x10<sup>-16</sup> | * |
| Coffee/Tobacco/Aspirin (3) | -0.7973 | 0.4506 (0.3832, 0.5298) | <2x10<sup>-16</sup> | * |
| Sex (Female) | 0.1172 | 1.1244 (1.0369, 1.2192) | 0.0045 | * |
| Study (Fox Insight) | 0.3555 | 1.4270 (1.1984, 1.6990) | 7x10<sup>-5</sup> | * |

coxph(formula = Surv(AAO/AAE, Diagnosis) ~ Coffee/Tobacco/Aspirin + Sex + Study, data = data);
\* p-value < 0.05
Abbreviations: AAO, age at onset; AAE, age at examination; PD, Parkinson's disease; CI, confidence interval.
Note: baseline categories: Coffee/Tobacco/Aspirin = 0, Sex = Male, Study = AMP-PD/PPMI.

We assessed the AAO ranges in the different groups of lifestyle factor exposures. In the subgroup of patients with PD that used no protective lifestyle factor, the median AAO was 57 years (range: 19-78 years), while patients with PD that drank coffee and used tobacco and aspirin had a median AAO of 66 years (range: 38-86 years), indicating a difference in the median AAO of 9 years in these two groups.

### Additive and interaction effects between PGS and lifestyle factors on AAO

Next, we explored the additive and interactive effects of the PGS and lifestyle factors on AAO in linear regression models. In the additive models including the lifestyle factor coffee drinking, the PGS was associated with AAO only when coffee drinking was used as a binary yes-no indication (β=-1.36, SE=0.69, p=0.0491) (Table 3). In addition, the positive association between the coffee drinking duration and AAO was still robust (β=0.18, SE=0.04, p=6x10^-5^). Further, there were no interactions between the PGS and coffee drinking in the patients with PD.

**Table 3.** Linear model on the association of PGS and coffee drinking with AAO in the PD study group.

|  | Estimate | Standard error | p-value |  |
| --- | --- | --- | --- | --- |
| **Coffee drinking (binary) (N=134)<sup>1</sup>** |  |  |  |  |
| Intercept | 57.0152 | 1.8829 | <2x10<sup>-16</sup> | * |
| PGS | -1.3647 | 0.6869 | 0.0491 | * |
| Coffee drinking (binary) | 1.6988 | 1.8102 | 0.3498 |  |
| Sex (Male) | 3.2507 | 1.6441 | 0.0502 |  |
| PC1 | -0.6476 | 74.6193 | 0.9931 |  |
| PC2 | -33.3712 | 55.7319 | 0.5504 |  |
| **Coffee drinking dosage (N=111)<sup>1</sup>** |  |  |  |  |
| Intercept | 57.5239 | 1.7765 | <2x10<sup>-16</sup> | * |
| PGS | -1.1348 | 0.7899 | 0.1538 |  |
| Coffee drinking dosage | 0.4448 | 0.6140 | 0.4705 |  |
| Sex (Male) | 1.8837 | 1.7584 | 0.2865 |  |
| PC1 | 38.1684 | 88.0026 | 0.6654 |  |
| PC2 | -65.5949 | 60.7299 | 0.2826 |  |
| **Coffee drinking duration (N=111)<sup>1</sup>** |  |  |  |  |
| Intercept | 54.8395 | 1.6555 | <2x10<sup>-16</sup> | * |
| PGS | -0.9684 | 0.7325 | 0.1890 |  |
| Coffee drinking duration | 0.1757 | 0.0421 | 6x10<sup>-5</sup> | * |
| Sex (Male) | 0.1953 | 1.6765 | 0.9075 |  |
| PC1 | 82.1193 | 81.7290 | 0.3173 |  |
| PC2 | -54.7535 | 56.4392 | 0.3342 |  |
| **Coffee drinking (binary) (N=135)<sup>2</sup>** |  |  |  |  |
| Intercept | 57.6190 | 2.0760 | <2x10<sup>-16</sup> | * |
| PGS | -1.3125 | 1.3469 | 0.3317 |  |
| Coffee drinking (binary) | 1.0096 | 2.2066 | 0.6481 |  |
| Sex (Male) | 3.5601 | 1.6658 | 0.0345 | * |
| PC1 | -51.0098 | 71.1716 | 0.4749 |  |
| PC2 | -28.6197 | 6.6410 | 0.6142 |  |
| PGS:Coffee drinking (binary) | -0.0375 | 1.5808 | 0.9811 |  |
| **Coffee drinking dosage (N=112)<sup>2</sup>** |  |  |  |  |
| Intercept | 58.4358 | 1.8485 | <2x10<sup>-16</sup> | * |
| PGS | -1.8856 | 1.1390 | 0.1008 |  |
| Coffee drinking dosage | -0.0688 | 0.6951 | 0.9213 |  |
| Sex (Male) | 2.2499 | 1.7801 | 0.2091 |  |
| PC1 | -32.1781 | 82.4732 | 0.6972 |  |
| PC2 | -52.4929 | 61.9505 | 0.3987 |  |
| PGS:Coffee drinking dosage | 0.6506 | 0.6813 | 0.3418 |  |
| **Coffee drinking duration (N=111)<sup>2</sup>** |  |  |  |  |
| Intercept | 55.0013 | 1.7524 | <2x10<sup>-16</sup> | * |
| PGS | -1.1931 | 1.0642 | 0.2648 |  |
| Coffee drinking duration | 0.1673 | 0.0512 | 0.0015 | * |
| Sex (Male) | 0.2092 | 1.6845 | 0.9014 |  |
| PC1 | 84.9419 | 82.6532 | 0.3065 |  |
| PC2 | -54.7779 | 56.6866 | 0.3361 |  |
| PGS:Coffee drinking duration | 0.0114 | 0.0391 | 0.7706 |  |

<sup>1</sup>glm(formula = AAO ~ PGS + Coffee drinking + Sex + PC1 + PC2, family = gaussian, data=data); \* p-value < 0.05
<sup>2</sup>glm(formula = AAO ~ PGS \* Coffee drinking + Sex + PC1 + PC2, family = gaussian, data= data); \* p-value < 0.05
Abbreviations: PGS, polygenic score; AAO, age at onset; PD, Parkinson's disease; PC, principal component.

When we investigated the association between PGS and tobacco use in an additive regression model, both the PGS (β=-1.11, SE=0.24, p=4x10^-6^) and tobacco use (binary ever-never indication) (β=3.21, SE=0.50, p=2x10^-10^) showed associations with AAO (Table 4). However, when dosage of tobacco was included (β=0.13, SE=0.08, p=0.1228), only the PGS (β=-1.49, SE=0.69, p=0.0337) was associated with AAO. Similarly, no association between the duration of tobacco use and AAO was found (β=0.12, SE=0.09, p=0.1781), while the association between the PGS and AAO (β=-1.56, SE=0.70, p=0.0275) was still robust. In addition, there were no interactions between the PGS and tobacco use with AAO. We further explored the association between the PGS and aspirin intake on AAO in an additive regression model. When aspirin intake was included as a binary yes-no indication, both the PGS (β=-1.63, SE=0.63, p=0.0112) and aspirin intake (β=7.61, SE=1.47, p=8x10^-7^) were associated with AAO in patients with PD (Table 5). Similarly, when the aspirin intake duration was included in the model, the PGS (β=-1.76, SE=0.69, p=0.0127), as well as aspirin intake duration (β=0.55, SE=0.15, p=0.0004), were associated with AAO. However, when including the aspirin intake dosage in the model, aspirin showed an association with AAO (β=0.79, SE=0.21, p=0.0003), while the association between PGS and AAO diminished (β=-1.12, SE=0.67, p=0.0664). There was further no interaction between PGS and aspirin intake in all interaction models.

**Table 4.**
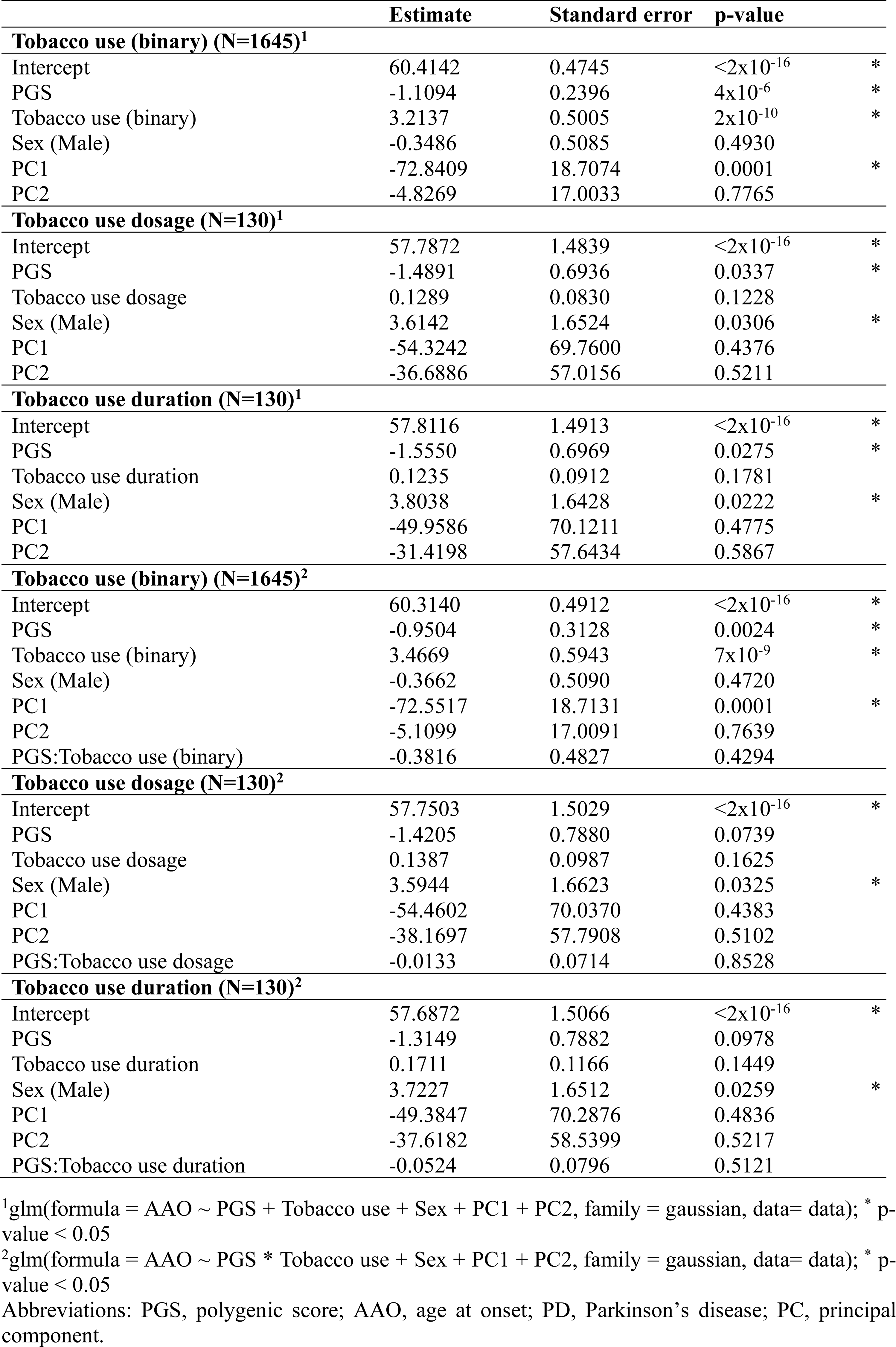
Linear model on the association of PGS and tobacco use with AAO in the PD study group.

|  | Estimate | Standard error | p-value |  |
| --- | --- | --- | --- | --- |
| **Tobacco use (binary) (N=1645)<sup>1</sup>** |  |  |  |  |
| Intercept | 60.4142 | 0.4745 | <2x10<sup>-16</sup> | * |
| PGS | -1.1094 | 0.2396 | 4x10<sup>-6</sup> | * |
| Tobacco use (binary) | 3.2137 | 0.5005 | 2x10<sup>-10</sup> | * |
| Sex (Male) | -0.3486 | 0.5085 | 0.4930 |  |
| PC1 | -72.8409 | 18.7074 | 0.0001 | * |
| PC2 | -4.8269 | 17.0033 | 0.7765 |  |
| **Tobacco use dosage (N=130)<sup>1</sup>** |  |  |  |  |
| Intercept | 57.7872 | 1.4839 | <2x10<sup>-16</sup> | * |
| PGS | -1.4891 | 0.6936 | 0.0337 | * |
| Tobacco use dosage | 0.1289 | 0.0830 | 0.1228 |  |
| Sex (Male) | 3.6142 | 1.6524 | 0.0306 | * |
| PC1 | -54.3242 | 69.7600 | 0.4376 |  |
| PC2 | -36.6886 | 57.0156 | 0.5211 |  |
| **Tobacco use duration (N=130)<sup>1</sup>** |  |  |  |  |
| Intercept | 57.8116 | 1.4913 | <2x10<sup>-16</sup> | * |
| PGS | -1.5550 | 0.6969 | 0.0275 | * |
| Tobacco use duration | 0.1235 | 0.0912 | 0.1781 |  |
| Sex (Male) | 3.8038 | 1.6428 | 0.0222 | * |
| PC1 | -49.9586 | 70.1211 | 0.4775 |  |
| PC2 | -31.4198 | 57.6434 | 0.5867 |  |
| **Tobacco use (binary) (N=1645)<sup>2</sup>** |  |  |  |  |
| Intercept | 60.3140 | 0.4912 | <2x10<sup>-16</sup> | * |
| PGS | -0.9504 | 0.3128 | 0.0024 | * |
| Tobacco use (binary) | 3.4669 | 0.5943 | 7x10<sup>-9</sup> | * |
| Sex (Male) | -0.3662 | 0.5090 | 0.4720 |  |
| PC1 | -72.5517 | 18.7131 | 0.0001 | * |
| PC2 | -5.1099 | 17.0091 | 0.7639 |  |
| PGS:Tobacco use (binary) | -0.3816 | 0.4827 | 0.4294 |  |
| **Tobacco use dosage (N=130)<sup>2</sup>** |  |  |  |  |
| Intercept | 57.7503 | 1.5029 | <2x10<sup>-16</sup> | * |
| PGS | -1.4205 | 0.7880 | 0.0739 |  |
| Tobacco use dosage | 0.1387 | 0.0987 | 0.1625 |  |
| Sex (Male) | 3.5944 | 1.6623 | 0.0325 | * |
| PC1 | -54.4602 | 70.0370 | 0.4383 |  |
| PC2 | -38.1697 | 57.7908 | 0.5102 |  |
| PGS:Tobacco use dosage | -0.0133 | 0.0714 | 0.8528 |  |
| **Tobacco use duration (N=130)<sup>2</sup>** |  |  |  |  |
| Intercept | 57.6872 | 1.5066 | <2x10<sup>-16</sup> | * |
| PGS | -1.3149 | 0.7882 | 0.0978 |  |
| Tobacco use duration | 0.1711 | 0.1166 | 0.1449 |  |
| Sex (Male) | 3.7227 | 1.6512 | 0.0259 | * |
| PC1 | -49.3847 | 70.2876 | 0.4836 |  |
| PC2 | -37.6182 | 58.5399 | 0.5217 |  |
| PGS:Tobacco use duration | -0.0524 | 0.0796 | 0.5121 |  |

<sup>1</sup>glm(formula = AAO ~ PGS + Tobacco use + Sex + PC1 + PC2, family = gaussian, data= data); \* p-value < 0.05
<sup>2</sup>glm(formula = AAO ~ PGS \* Tobacco use + Sex + PC1 + PC2, family = gaussian, data= data); \* p-value < 0.05
Abbreviations: PGS, polygenic score; AAO, age at onset; PD, Parkinson's disease; PC, principal component.

**Table 5.** Linear model on the association of PGS and aspirin intake with AAO in the PD study group.

|  | Estimate | Standard error | p-value |  |
| --- | --- | --- | --- | --- |
| **Aspirin intake (binary) (N=135)<sup>1</sup>** |  |  |  |  |
| Intercept | 56.2154 | 1.3937 | <2x10<sup>-16</sup> | * |
| PGS | -1.6302 | 0.6332 | 0.0112 | * |
| Aspirin intake (binary) | 7.6120 | 1.4665 | 8x10<sup>-7</sup> | * |
| Sex (Male) | 2.5009 | 1.4978 | 0.0974 |  |
| PC1 | -35.7824 | 63.9946 | 0.5770 |  |
| PC2 | -39.8831 | 51.0860 | 0.4364 |  |
| **Aspirin intake dosage (N=129)<sup>1</sup>** |  |  |  |  |
| Intercept | 57.3304 | 1.5113 | <2x10<sup>-16</sup> | * |
| PGS | -1.2413 | 0.6703 | 0.0664 |  |
| Aspirin intake dosage | 0.7876 | 0.2102 | 0.0003 | * |
| Sex (Male) | 2.4337 | 1.6244 | 0.1366 |  |
| PC1 | -56.1517 | 67.9612 | 0.4103 |  |
| PC2 | -41.6697 | 54.3264 | 0.4445 |  |
| **Aspirin intake duration (N=113)<sup>1</sup>** |  |  |  |  |
| Intercept | 56.4635 | 1.5251 | <2x10<sup>-16</sup> | * |
| PGS | -1.7575 | 0.6930 | 0.0127 | * |
| Aspirin intake duration | 0.5455 | 0.1485 | 0.0004 | * |
| Sex (Male) | 3.4999 | 1.6937 | 0.0412 | * |
| PC1 | 0.6326 | 73.6632 | 0.9932 |  |
| PC2 | 10.3268 | 56.1647 | 0.8545 |  |
| **Aspirin intake (binary) (N=135)<sup>2</sup>** |  |  |  |  |
| Intercept | 56.2268 | 1.4639 | <2x10<sup>-16</sup> | * |
| PGS | -1.6447 | 0.8406 | 0.0526 |  |
| Aspirin intake (binary) | 7.5860 | 1.7713 | 4x10<sup>-5</sup> | * |
| Sex (Male) | 2.4994 | 1.5047 | 0.0991 |  |
| PC1 | -35.9680 | 64.6249 | 0.5788 |  |
| PC2 | -39.8840 | 51.2850 | 0.4382 |  |
| PGS:Aspirin intake (binary) | 0.0341 | 1.2896 | 0.9789 |  |
| **Aspirin intake dosage (N=129)<sup>2</sup>** |  |  |  |  |
| Intercept | 57.3660 | 1.5761 | <2x10<sup>-16</sup> | * |
| PGS | -1.2812 | 0.8252 | 0.1231 |  |
| Aspirin intake dosage | 0.7779 | 0.2401 | 0.0016 | * |
| Sex (Male) | 2.4217 | 1.6373 | 0.1417 |  |
| PC1 | -56.6835 | 68.5336 | 0.4098 |  |
| PC2 | -42.0503 | 54.7370 | 0.4438 |  |
| PGS: Aspirin intake dosage | 0.0161 | 0.1924 | 0.9336 |  |
| **Aspirin intake duration (N=112)<sup>2</sup>** |  |  |  |  |
| Intercept | 55.9855 | 1.5538 | <2x10<sup>-16</sup> | * |
| PGS | -1.5249 | 0.7712 | 0.0506 |  |
| Aspirin intake duration | 0.8297 | 0.2151 | 0.0002 | * |
| Sex (Male) | 3.6622 | 1.6844 | 0.0319 | * |
| PC1 | 17.4621 | 73.4965 | 0.8127 |  |
| PC2 | 32.5304 | 57.5683 | 0.5732 |  |
| PGS:Aspirin intake duration | -0.2382 | 0.1793 | 0.1869 |  |

<sup>1</sup>glm(formula = AAO ~ PGS + Aspirin intake + Sex + PC1 + PC2, family = gaussian, data= data); \* p-value < 0.05
<sup>2</sup>glm(formula = AAO ~ PGS \* Aspirin intake + Sex + PC1 + PC2, family = gaussian, data= data); \* p-value < 0.05
Abbreviations: PGS, polygenic score; AAO, age at onset; PD, Parkinson's disease; PC, principal component.

### Impact of PGS and lifestyle factors on AAO

Since the association between PGS and AAO diminished in some models including lifestyle factors as covariables, we investigated the impact PGS and lifestyle factors have on the AAO in PD. In a first approach to compare the effect sizes of PGS and lifestyle factors on AAO, we categorized the PGS into “low PGS” and “high PGS” according to the median PGS and stratified participants into the subgroups that either used no protective lifestyle factor or that used all three lifestyle factors. In the subgroup of participants that used no protective lifestyle factor, a high PGS showed a 3.03 times higher expected hazard of PD as compared to a low PGS (HR=3.03, 95% CI=1.05-8.78, p=0.0409). In contrast, in the subgroup of participants that used all three lifestyle factors, there was no increased hazard ratio (HR=1.24, 95% CI=0.48-3.19, p=0.6577).

We further investigated the model goodness-of-fit of the linear models using the adjusted deviance-based R^2^. The model assessing the association between PGS and AAO, while using sex and the first two PCs as covariables, had an adjusted R^2^ of 0.0141. In contrast, the linear model evaluating the association between the three lifestyle factors and AAO with the same covariables had an adjusted R^2^ of 0.1079, when the lifestyle factors were coded as cumulative quantitative numbers.

### Relationship between lifestyle factors and AAO in GBA1-PD

To investigate if the individual and combined effects of the lifestyle factors coffee, tobacco, and aspirin are exclusive to idiopathic PD or if these effects can also be found in patients who carry *GBA1* variants, which are considered some of the strongest genetic risk variants for PD, we examined the relationship between the protective lifestyle factors and AAO in an additional study group of patients with *GBA1*-PD from Fox Insight. In the linear regression model, coffee drinking duration was positively associated with AAO (β=0.48, SE=0.05, p=1x10^-12^) in *GBA1*-PD. In addition, we observed a positive association between tobacco use and AAO (β=3.65, SE=1.58, p=0.0223). Interestingly, aspirin intake did not show any associations with AAO, which contrasts with the results found in idiopathic PD. When including all three lifestyle factors separately in the same model to predict AAO, only tobacco use was associated with AAO (β=6.78, SE=2.17, p=0.0024). To examine the combined effect of the three lifestyle factors in more detail, we used as influence variable the cumulative number of factors in the Cox proportional hazards model. We observed a protective trend for this variable. With no lifestyle factor as reference, we observed an HR of 0.85 for the use of one (95% CI=0.40-1.78, p=0.6648), HR of 0.47 for the use of two (95% CI=0.23-0.97, p=0.0410), and an HR of 0.35 for the use of three lifestyle factors (95% CI=0.14-0.86, p=0.0216). Of note, the use of one lifestyle factor did not show a significant reduction in the hazard ratio compared to the use of none of these lifestyle factors, which could be a problem of statistical power. However, the use of two or three lifestyle factors showed a significant reduction in the hazard ratio by 53% and 65%, indicating a protective effect on the AAO when using more lifestyle factors.

## Discussion

In this study, we have investigated the association between the PGS, calculated based on a previously proposed composition of 1805 variants^2^, and the AAO in patients with PD and determined the interaction between the PGS and the lifestyle factors coffee drinking, tobacco use, and aspirin intake on the AAO in PD.

We found that the PGS not only allows discrimination between PD cases and controls^2,4^ but also showed a negative correlation with AAO^4,10,12^, indicating that an increase of the PGS by one SD leads to an approximately one year earlier AAO in patients with PD. This relationship between PGS and AAO was also robust when adjusting for potentially confounding covariables (i.e., sex and ancestry as represented by the first two principal components). These results demonstrate that the genetic composition, represented by the PGS, adds to understanding the variance in AAO in patients with PD. However, with a range in AAO of more than 70 years in this study group, more influencing factors and cofounders need to be considered.

The protective effect of environmental and lifestyle factors that decrease the risk of developing PD, influence initial PD-related symptoms and progression, and delay AAO has already been known for years^40,41^. However, how these lifestyle factors interact and which combined effect they have on PD AAO remains unresolved. Our group has previously presented a protective effect of coffee, tobacco, and aspirin on the AAO of patients with PD from the Fox Insight study^31^, which we further replicated in the AMP-PD/PPMI study group here. This correlation between lifestyle factors and PD AAO consistently highlights the importance of investigating this interplay further. In a more detailed analysis of the combined effect of the three protective lifestyle factors coffee, tobacco, and aspirin on AAO, we found that the use of either one, two, or three lifestyle factors led to a reduction in the hazard ratio by 24%, 46%, and 59%, respectively, in comparison to no use. These hazard ratio values are consistent with additive, i.e., independent effects on the logit scale of the lifestyle factors with no synergistic interaction, indicating different underlying mechanisms that lead to the later AAO. Deciphering these mechanisms of action is important to develop suitable therapeutic strategies to delay the AAO of patients with PD. Interestingly, aspirin seems to have a larger effect on AAO than coffee or tobacco. Given that inflammation is a crucial pathophysiological pathway in PD^42^, the anti-inflammatory effect of aspirin might have a protective impact on PD AAO. Although a positive effect of non-steroidal anti-inflammatory drugs on PD risk is contentious^43^, it is well-known that sustained neuroinflammation leads to the progressive degeneration of dopaminergic neurons^44^. The intake of anti-inflammatory drugs such as aspirin in the prodromal phase, when neuronal degeneration has already started, might therefore slow this process resulting in a later AAO. Interestingly, there was no protective effect of aspirin on the AAO of patients with *GBA1*-PD, indicating that the neuroinflammatory mechanisms leading to neurodegeneration might diverge from patients with idiopathic PD or be masked by the genetic susceptibility in *GBA1*-PD. Although the pro-inflammatory signaling does not seem to be related to the PD subtype and there is no evidence of a difference in the immune response between idiopathic PD, monogenic forms of PD (e.g., *LRRK2*-PD), and strong risk factor carriers such as *GBA1*-PD^45^, those PD subtypes present with different phenotypes and might need to be treated differently. Thus far, it is not clear how the underlying mechanisms work and if they differ in the different subtypes of PD. However, we have already seen that the effects of environmental and lifestyle factors as well as specific genetic risk factors on AAO vary in different subtypes of PD, especially between monogenic forms of PD and idiopathic PD^46^. To follow up on this, future larger-scale studies including patients with monogenic forms of PD or who carry strong risk factors (e.g., *LRRK2*-PD, *GBA1*-PD, or *PINK1/Parkin*-PD), are important to target the effect of anti-inflammatory lifestyle factors on PD AAO.

In order to investigate the additive and interaction effects of the lifestyle factors together with the PGS on AAO, we applied linear models, showing additive and independent effects of PGS and tobacco use on AAO as well as of PGS and aspirin intake on AAO, with opposite directionality of the PGS and the lifestyle factors. The possibility of an interaction between PGS and lifestyle factors cannot be entirely ruled out. We found a three times higher expected hazard of PD in the subgroup of participants that used no protective lifestyle factor and had a high PGS as compared to patients with a low PGS. In contrast, in the subgroup of participants that used all three lifestyle factors, there was no increased hazard of PD between a high and a low PGS. However, the sample sizes in these models were low with only n=21 patients with PD and confidence intervals were large. Therefore, the results are thus far only preliminary and need to be interpreted with caution. Nevertheless, they indicate that the PGS is more important for persons that use no protective lifestyle factors and that PGS and lifestyle factors could at least in part cover the same effect. We also found the three lifestyle factors to explain the AAO in patients with PD more accurately than the PGS (Lifestyle factor model: R^2^=0.1079, PGS model: R^2^=0.0141).

Although there are known gene-environment interactions of coffee and tobacco^20–23^ with PD, none of the variants included in the calculation of the PGS are located within genes known to show interactions with coffee, smoking, or aspirin. Since this PGS is based on common variants associated with PD risk, it is pathway-independent and different mechanisms can lead to the earlier AAO when having a high PGS or the delayed AAO when using protective lifestyle factors. In contrast, pathway-dependent PGSs such as the mitochondrial polygenic score^46,47^ have been shown to interact with lifestyle factors such as pesticides or caffeinated beverages.

Limitations of our study include clinical and genetic data harmonization. The use of data from different cohorts poses the problem of overcoming potential inconsistencies due to differences in the way of data collection. To help overcome this problem, we corrected for the study site in our Cox proportional hazards models including lifestyle data from AMP-PD/PPMI and Fox Insight and also corrected for the first two principal components in all genetic data analyses to account for genetic differences due to ethnic diversity or differences in the type of genetic data collection. Another limitation was that the clinical data provided by the three cohorts sparsely overlapped with genetic data, resulting in small sample sizes in some of the subgroups. Nevertheless, we showed that lifestyle factors have an important effect on PD AAO that is even greater than that of a combined genetic risk. In future studies, this analysis needs to be replicated in a larger study group with diverse ancestral backgrounds. Since the GWAS that was used for the PGS calculation was performed in a European ancestry population, we only included PD patients and controls with European ancestry in our study group. The lack of ancestry and ethnic diversity in large-scale genetic studies is a well-known problem^48–50^. To completely unravel the genetic mechanisms that lead to developing PD, future studies must be inclusive of patients from all cultural and genetic backgrounds.

In conclusion, this study is the first to assess the combined effect of the PD-specific PGS together with coffee drinking, tobacco use, and aspirin intake on the AAO of patients with PD and adds to understanding this complex disease. Our results further indicate a potential neuroprotective role of the anti-inflammatory drug aspirin resulting in a later AAO in PD. Aspirin might play an important protective part in the inflammatory processes that could lead to neurodegeneration in PD. Thus far, these results are only exploratory, because they did not follow an “a priori” hypothesis and further validation is essential. Nevertheless, our findings underline the importance of investigating both genetic disposition and external influences such as environmental and lifestyle factors to unravel the likelihood of disease manifestation and the variable phenotype presented in patients with PD.

## Data Availability

All data produced in the present study are available upon reasonable request to the authors and with permission of the Accelerating Medicine Partnership Parkinson's Disease Knowledge Platform and the Michael J. Fox Foundation.

## Declarations

## Acknowledgments

A special thanks to the families and patients who participated in this study.

We would also like to thank Mike Nalls for providing the list of the 1805 common risk variants including the reference alleles and effect sizes that were used in the PGS calculation.

This study was funded by The Michael J. Fox Foundation for Parkinson’s Research (MJFF-019271 and MJFF-021227). This project was further supported by the Institute of Neurogenetics at the University of Lübeck. Grant support was received from the DFG RU ProtectMove (FOR2488).

The AMP PD program is a public-private partnership managed by the Foundation for the National Institutes of Health and funded by the National Institute of Neurological Disorders and Stroke (NINDS) in partnership with the Aligning Science Across Parkinson’s (ASAP) initiative; Celgene Corporation, a subsidiary of Bristol-Myers Squibb Company; GlaxoSmithKline plc (GSK); The Michael J. Fox Foundation for Parkinson’s Research; Pfizer Inc.; Sanofi US Services Inc.; and Verily Life Sciences. Accelerating Medicines Partnership and AMP are registered service marks of the U.S. Department of Health and Human Services. Clinical data and biosamples used in preparation of this article were obtained from the (i) Michael J. Fox Foundation for Parkinson’s Research (MJFF) and National Institutes of Neurological Disorders and Stroke (NINDS) BioFIND study, (ii) Harvard Biomarkers Study (HBS), (iii) National Institute on Aging (NIA) International Lewy Body Dementia Genetics Consortium Genome Sequencing in Lewy Body Dementia Case-control Cohort (LBD), (iv) MJFF LRRK2 Cohort Consortium (LCC), (v) NINDS Parkinson’s Disease Biomarkers Program (PDBP), (vi) MJFF Parkinson’s Progression Markers Initiative (PPMI), and (vii) NINDS Study of Isradipine as a Disease-modifying Agent in Subjects With Early Parkinson Disease, Phase 3 (STEADY-PD3) and (viii) the NINDS Study of Urate Elevation in Parkinson’s Disease, Phase 3 (SURE-PD3). BioFIND is sponsored by The Michael J. Fox Foundation for Parkinson’s Research (MJFF) with support from the National Institute for Neurological Disorders and Stroke (NINDS). The BioFIND Investigators have not participated in reviewing the data analysis or content of the manuscript. For up-to-date information on the study visit michaeljfox.org/news/biofind. Genome sequence data for the Lewy body dementia case-control cohort were generated at the Intramural Research Program of the U.S. National Institutes of Health. The study was supported in part by the National Institute on Aging (program #: 1ZIAAG000935) and the National Institute of Neurological Disorders and Stroke (program #: 1ZIANS003154). The Harvard Biomarker Study (HBS) is a collaboration of HBS investigators (full list of HBS investigators at https://www.bwhparkinsoncenter.org/biobank/) and funded through philanthropy and NIH and Non-NIH funding sources. The HBS Investigators have not participated in reviewing the data analysis or content of the manuscript. PPMI is sponsored by The Michael J. Fox Foundation for Parkinson’s Research and supported by a consortium of scientific partners (full list of all of the PPMI funding partners found at https://www.ppmi-info.org/about-ppmi/who-we-are/study-sponsors). The PPMI investigators have not participated in reviewing the data analysis or content of the manuscript. For up-to-date information on the study, visit www.ppmi-info.org. The Parkinson’s Disease Biomarker Program (PDBP) consortium is supported by the National Institute of Neurological Disorders and Stroke (NINDS) at the National Institutes of Health. A full list of PDBP investigators can be found at https://pdbp.ninds.nih.gov/policy. The PDBP investigators have not participated in reviewing the data analysis or content of the manuscript. The Study of Isradipine as a Disease-modifying Agent in Subjects With Early Parkinson Disease, Phase 3 (STEADY-PD3) is funded by the National Institute of Neurological Disorders and Stroke (NINDS) at the National Institutes of Health with support from The Michael J. Fox Foundation and the Parkinson Study Group. For additional study information, visit https://clinicaltrials.gov/ct2/show/study/NCT02168842. The STEADY-PD3 investigators have not participated in reviewing the data analysis or content of the manuscript. The Study of Urate Elevation in Parkinson’s Disease, Phase 3 (SURE-PD3) is funded by the National Institute of Neurological Disorders and Stroke (NINDS) at the National Institutes of Health with support from The Michael J. Fox Foundation and the Parkinson Study Group. For additional study information, visit https://clinicaltrials.gov/ct2/show/NCT02642393. The SURE-PD3 investigators have not participated in reviewing the data analysis or content of the manuscript.

## Author Contributions

CG performed the data analysis, data interpretation, and writing of the manuscript. LB performed the data analysis, and data interpretation and reviewed the manuscript. TL contributed to the data analysis and interpretation and reviewed the manuscript. IRK helped with the data interpretation and reviewed the manuscript. AC helped with the data interpretation and reviewed the manuscript. SK helped with the data analysis, and data interpretation, and reviewed the manuscript. BHL helped with the data interpretation and reviewed the manuscript. CK helped with the data interpretation and reviewed the manuscript. JT was responsible for the conceptualization and design of the project, data interpretation, and writing of the manuscript.

## Data availability

Data used in the preparation of this article were obtained from the Accelerating Medicine Partnership® (AMP®) Parkinson’s Disease (AMP PD) Knowledge Platform. For up-to-date information on the study, visit https://www.amp-pd.org. The data that support the findings of this study are available from the Accelerating Medicine Partnership® (AMP®) Parkinson’s Disease Knowledge Platform, but restrictions apply to the availability of these data, which were used under license for the current study, and so are not publicly available. Data are however available from the authors upon reasonable request and with permission of the Accelerating Medicine Partnership® (AMP®) Parkinson’s Disease Knowledge Platform.

Data used in the preparation of this manuscript were obtained from the Fox Insight database (https://foxinsight-info. michaeljfox.org/insight/explore/insight.jsp) on 22/05/2023. For up-to-date information on the study, visit https://foxinsight-info.michaeljfox.org/ insight/explore/insight.jsp. The data that support the findings of this study are available from the Fox Insight Study, sponsored by the Michael J. Fox Foundation, but restrictions apply to the availability of these data, which were used under license for the current study, and so are not publicly available. Data are however available from the authors upon reasonable request and with permission of the Michael J. Fox Foundation.

Data used in the preparation of this article were obtained from the Parkinson’s Progression Markers Initiative (PPMI) database (www.ppmi-info.org/access-data-specimens/download-data), RRID:SCR_006431. For up-to-date information on the study, visit www.ppmi-info.org.

